# Early insights into nurses’ perceptions, opportunities, and concerns regarding Ambient artificial intelligence in nursing practice

**DOI:** 10.64898/2026.09.02.26362036

**Authors:** Yu-Ching Yang, Meghan Reading Turchioe, Afra Shamnath, Sarah C. Rossetti

## Abstract

Documentation burden remains a persistent challenge in nursing practice, and Ambient artificial intelligence (Ambient AI) may offer new opportunities to mitigate the burden and improve nursing practice. This brief scientific communication reports survey findings on the current status of Ambient AI implementation in U.S. inpatient nursing settings and nurses’ perceptions of its anticipated benefits and concerns. We conducted a cross-sectional online survey of practicing inpatient nurses recruited from nursing-focused social media communities. Descriptive analyses were used to summarize Ambient adoption, user experience, and perceived benefits and concerns. Among 61 eligible respondents, nearly 60% (n = 37) reported that Ambient AI had been piloted or implemented in nursing at their healthcare institutions, and approximately half (49.2 %, n = 30) reported having direct experience using Ambient AI in clinical practice. Most nurses with direct experience (73.3%, n = 22) described positive experience using the technology in practice. The frequently identified perceived benefits of Ambient AI include reduced documentation time and improved patient care workflow, and improved data quality. However, nurses also raised concerns about patients’ acceptance of the new technology in their care, potential nursing job displacement, and loss of nursing skills due to overreliance on AI technology. Findings suggest that Ambient AI may help reduce nursing documentation burden and workload, yet implementation strategies should address nurses’ concerns about its influence on nursing professions, foster trust among clinicians and patients, and support integration into everyday nursing practice.

## Introduction

Documentation burden remains a persistent challenge in contemporary healthcare settings and has important implications for nurses’ workload, care quality, and patient safety. Although nursing documentation serves essential clinical and legal functions (Ho et al., 2014; Kuhn et al., 2015), it requires substantial time and attention during daily practice. Previous studies have shown that nurses spend nearly 23% of their daily working time on documentation-related activities (Baumann et al., 2018; Moore et al., 2020). These time demands are accompanied by significant cognitive and workflow burden, as nurses must constantly shift attention between documenting clinical information and managing ongoing patient care responsibilities (Harris et al., 2018; Melnick et al., 2021; Shan et al., 2023). Competing demands on nurses’ time and fragmented workflows reduce the time available for direct patient care, with potential negative consequences for the quality of care and patient safety (Shan et al., 2023). Although strategies such as standardized electronic templates and mobile documentation systems have improved documentation efficiency (Ehrler et al., 2021); they primarily streamline the process rather than reducing the overall workload associated with documentation. These persistent documentation-related burdens highlight the need for innovative approaches that reduce time in documentation while preserving the accuracy, completeness, and essential role of documentation in supporting patient care.

Recent advances in Ambient artificial technology, or Ambient AI, have created new opportunities to address documentation challenges in healthcare. Ambient AI refers to AI systems embedded within a clinical environment that unobtrusively collect, process, and interpret information from routine care activities to support clinical work and decision-making (Guo et al., 2026). One promising application is clinical documentation support. Unlike traditional electronic health record documentation, which requires clinicians to manually enter information, ambient AI operates in the background of the clinical workflow by capturing clinician-patient conversations and transforming them into draft clinical notes using speech recognition and natural language processing technologies (Guo et al., 2026). By integrating documentation into the natural workflow of care, Ambient AI has the potential to reduce manual documentation time and minimize workflow interruptions, and allow nurses to devote more time to direct, patient-centered care.

Although Ambient AI remains an emerging technology, early implementation efforts have been reported across a variety of clinical settings (Nahar & Kachnowski, 2023). Findings from these pilot studies suggest that Ambient AI may reduce physicians’ documentation time (Duggan et al., 2025; Lukac et al., 2025; Stults et al., 2025) and enhance user satisfaction (Duggan et al., 2025; Stults et al., 2025). However, existing evidence has largely focused on physicians’ perspectives and use cases (Razaghi et al., 2026). Little is known about the implementation of Ambient AI in nursing practice and nurses’ perceptions of its potential benefits and challenges. This gap is important as nursing documentation differs substantially from physicians’ documentation in its scope, workflow, and integration into continuous patient care. In addition, nurses represent the largest professional group in the healthcare workforce and account for the highest volume of data entered in the hospital setting; as such, in the hospital setting, nurses have the potential to be primary users of Ambient technology. Hence, understanding their perspectives is critical for successful integration into clinical practice. The purpose of the study is to examine the current status of Ambient AI implementation in nursing practice and to explore practicing nurses’ perceptions of anticipated benefits and concerns. We focus on inpatient practicing nurses because they manage extensive documentation requirements within a highly dynamic clinical environment that may be significantly affected by Ambient AI-enabled documentation support. Findings from the study may provide insights into nurses’ perspectives on Ambient AI and inform future research on its implementation strategies for integrating Ambient AI into nursing practice.

## 2. Methods

We conducted a cross-sectional anonymous online survey of practicing nurses recruited through social media platforms. This study was approved by the Institutional Review Board at our institution. Eligible participants were (1) self-identified as registered nurses or licensed practicing nurses and were actively employed in a U.S. healthcare institution, and (2) reported working in inpatient clinical settings, including the emergency department, floor and step-down unit, critical care, and operating room, at the time of survey participation. Participants were recruited using convenience sampling through nursing-focused communities on Facebook and LinkedIn. Online recruitment announcements were posted to approximately 55 nursing-focused communities on Facebook and LinkedIn and reposted periodically during the study period. Interested nurses could access the survey through a Qualtrics link. The survey included 17 items assessing participants’ demographics and professional characteristics, experiences with Ambient adoption, and perceived benefits and concerns. After survey completion, participants had the option to enter a raffle drawing for one of three $100 gift cards. Raffle contact information was collected separately from survey responses to maintain anonymity. No personally identifiable information or IP addresses were collected. Qualtrics security features, including bot detection and human verification, were enabled to reduce automated responses.

Standard data cleaning approaches were used to identify and remove automated responses, including low reCAPTCHA score (< 0.50) or demonstrated suspicious response patterns indicative of automated activity, such as duplicate responses or nonsensical, repetitive, or irrelevant open-ended content. A total of 119 human-generated survey responses were received between April and June 2026. Of these, 58 from individuals who self-identified as advanced practice registered nurses or nurse leaders were excluded, as the study focused on practicing inpatient nurses. The final analytic sample included 61 eligible inpatient nurses. Descriptive analyses were conducted to summarize participants’ characteristics and user experience of ambient AI. A large language model (ChatGPT Education, Version 5.5) supported data management, variable recoding, and generation of descriptive findings using structured prompts. All AI-assisted procedures complied with our institutional guidelines and training on AI-assisted research and educational applications. A research team member (YY) verified all analysis results using Microsoft Excel.

## 3. Results

The final analytic sample included 61 practicing inpatient nurses from 27 U.S. states. Respondents most commonly resided in the South (39.3%, n= 24), followed by the West (26.2%, n = 16), Midwest (24.6, n = 15), and Northeast (8.2%, n = 5). California accounted for the largest proportion of respondents (21.3%, n = 13). Most respondents were between 25 and 44 years of age (93.4%, n = 57), female (57.4%, n = 35), and held a bachelor’s degree (62.3%, n = 38). Nearly 70% (n = 42) reported 3 – 10 years of nursing experience. Participants represented diverse healthcare organizations, including community-based healthcare systems (37.5%, n = 39), academic medical centers (32.8%, n = 20). Respondents also represented a wide range of nursing specialties, primarily emergency and critical care nursing (55.8%, n = 34) and nurse anesthetists (14.8%, n = 9).

Most respondents reported organizational exposure to Ambient AI. Nearly 80% (n=48) indicated that Ambient AI had been piloted or implemented within their institutions (Table 1). Approximately 60% (n=37) reported that implementation involved registered nurses, either exclusively (36.0%, n = 22) or in combination with other healthcare professionals (20.3%, n = 15). Of 61 respondents, about half (49.2 %, n = 30) reported direct experience using Ambient AI in clinical practice. Reported use occurred most commonly in non-intensive care inpatient units (27.9%, n =17), emergency departments (18.0%, n = 11), intensive care units (16.4%, n = 10), or operating rooms (14.8%, n = 9). Among respondents with direct experience using Ambient AI, 73.3% (n = 22) expressed an overall positive experience, while 26.7% (n = 8) described a mix of positive and negative experiences.

**Table 1.** Current status of Ambient AI pilot or implementation in U.S. healthcare organizations.

| Ambient AI implementation group | n | % |
| --- | --- | --- |
| With registered nurses | 22 | 35.9 |
| Registered nurses and other health professionals | 15 | 23.4 |
| With other healthcare professionals | 13 | 20.3 |
| No/Unsure | 11 | 20.3 |

Overall, of 61 practicing nurses, 75.4% (n = 46) reported enthusiasm about the potential of Ambient AI to improve nursing practice. The most frequently identified potential benefits were reduced documentation time (93.4%, n = 57), improved patient workflow (86.9%, n = 53), and improved clinical data quality (86.9%, n = 53) (Figure 1). Qualitative comments from nurses with direct experience noted that the Ambient AI technology made documentation “very fast and easier”, reduced “time spent on manual charting,” and helped “save a lot of time on documentation during busy ED shifts.” In contrast, improved communication (75.4%, n = 46) and patient safety (70.5%, n = 43) were among the least frequently perceived potential benefits. Despite these positive perceptions, nurses also identified concerns about the potential impact of AI on the nursing profession. The most frequently reported concerns were patient distrust in AI-supported care (52.5%, n = 32), followed by loss of clinical skills due to reliance on AI (45%, n = 55), nursing job displacement (38%, n = 47), and increased nursing workload related to errors or correction (36.1%, n= 22) (Figure 2). Among nurses with direct experience using Ambient AI, additional concerns centered on documentation accuracy and privacy issues associated with continuous listening technologies.

**Figure 1.**
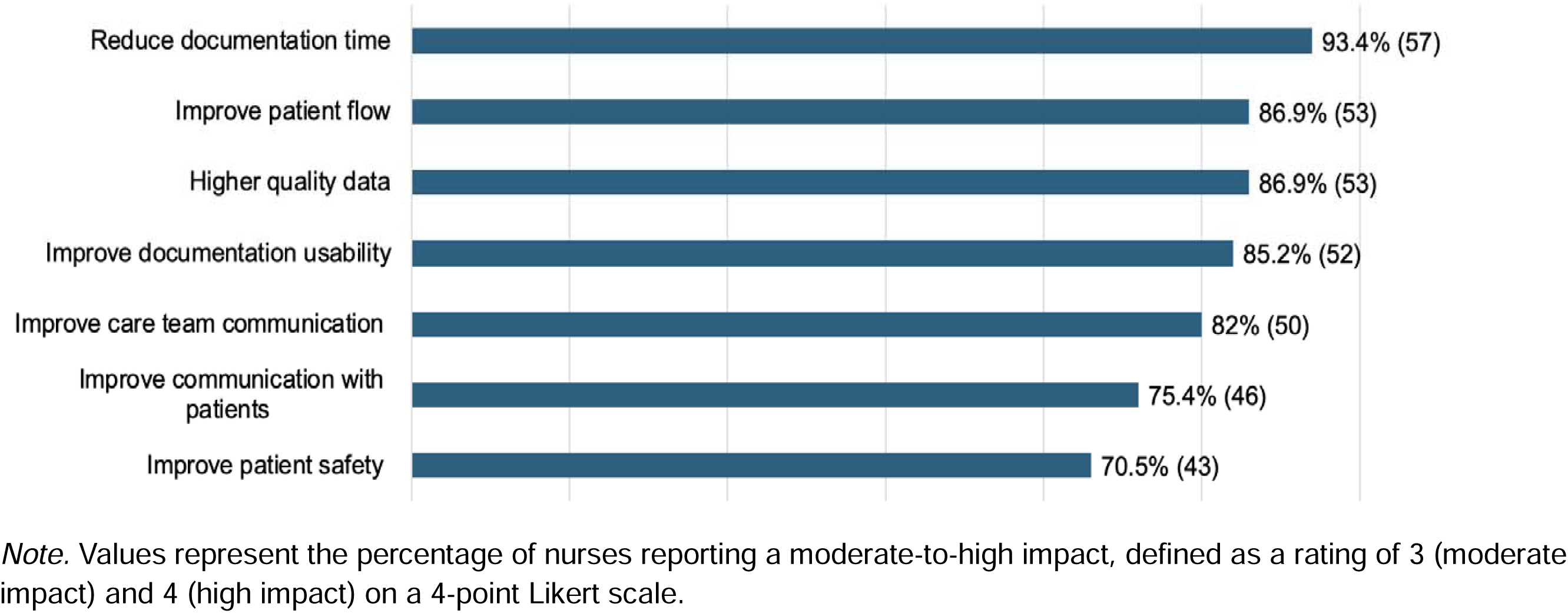
Perceived potential positive impact of Ambient AI in nursing practice (N=61)

**Figure 2.**
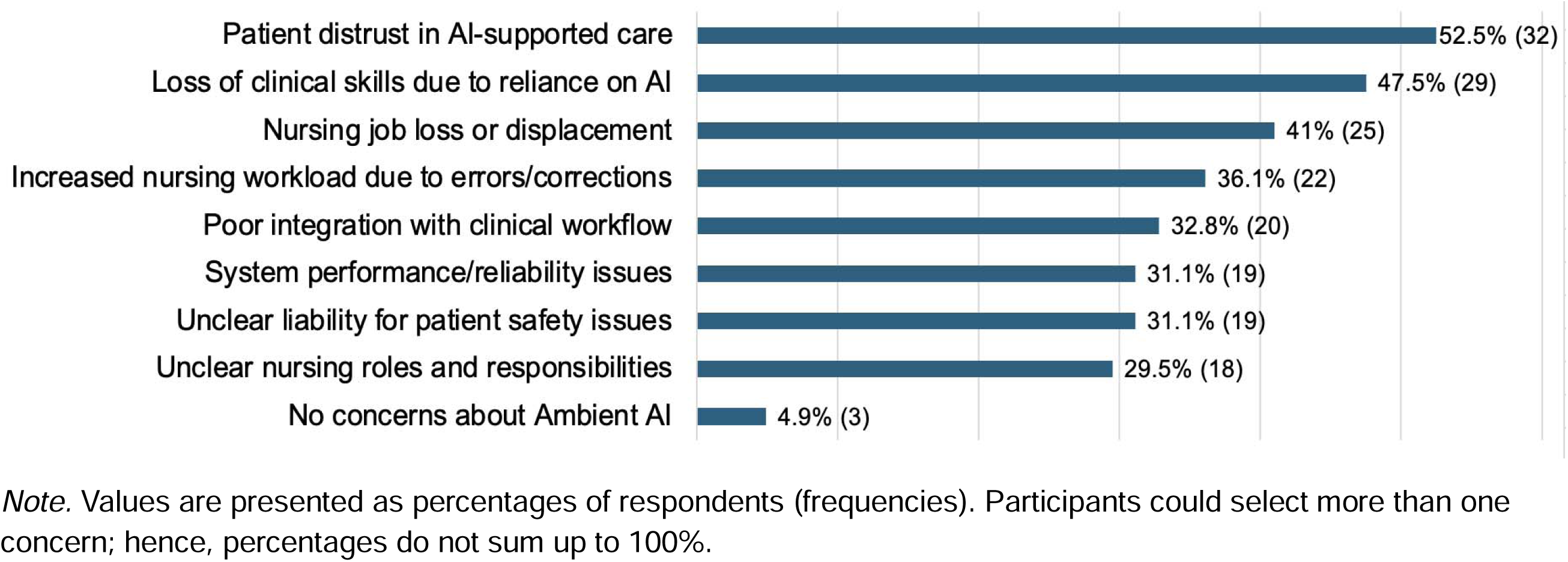
Nurses’ concerns about Ambient AI in nursing practice (N=61)

## 4. Discussion

This brief report provides early insights into Ambient AI implementation in nursing practice and nurses’ perceptions of its anticipated impact on patient care and professional practice. Overall, findings suggest that Ambient AI is increasingly being piloted or implemented across some U.S. healthcare institutions and is generally viewed positively by nurses with direct experience using the technology. The most frequently perceived potential benefits were reduced documentation time and improved patient care workflow, suggesting that Ambient AI may help mitigate nurses’ documentation-related workload and cognitive burden among practicing nurses. Despite these positive perceptions, perceived improvement in patient safety and nurse-patient communication were reported less frequently, and many nurses questioned whether patients would accept AI-enabled care. Nurses also raised concerns about loss of clinical skills due to overreliance on AI and potential nursing job displacement. These findings suggest that nurses recognize Ambient AI’s potential to reduce clinical workload but remain unclear about its value for patient-centered care and its long-term implications for the nursing profession. Future implementation efforts should emphasize that ambient AI supports, rather than replaces, nurses by reducing documentation time and burden. By easing the documentation burden, Ambient AI may allow nurses to devote more time and clinical attention to patient-centered care, where nursing expertise, judgement, and communication are most essential.

The study has several limitations. First, the study used convenience sampling through social media, which may have introduced selection bias. Nurses who responded to the survey may have been more interested in Ambient AI than the broader nursing workforce. Findings may not be fully representative of all U.S. nurses’ perspectives. Second, recruitment through social media posed data quality challenges related to potential fraudulent or automated survey responses. Although multiple screening procedures were applied to identify and remove suspicious responses, these challenges reduced the number of eligible responses and may have affected the final analytic sample. Finally, the final analytic sample was relatively small and limited to practicing nurses working in inpatient settings, which may limit generalizability of findings to outpatients, home care, or other clinical environments.

## 5. Conclusion

Ambient AI is emerging as a promising technology with the potential to reduce documentation burden and improve workflow efficiency in nursing practice. While nurses appear enthusiastic abouts it clinical application, its successful implementation requires strategies that build nurses’ trust and address concerns about its potential implications for the nursing profession and patient care.

## 6. Human subjects

The study was performed in compliance with the World Medical Association Declaration of Helsinki on Ethical Principles for Medical Research Involving Human Subjects, and was reviewed by the Columbia University Institutional Review Board.

## Declaration of generative AI and AI-assisted technologies in the writing process

During the preparation, the first author (YY) drafted the initial manuscript and used ChatGPT primarily to correct typos and grammar and to improve academic word choice. The author takes full responsibility for the final version of the manuscript.

## Data Availability

All data produced in the present study are available upon reasonable request to the authors

